# Functional Profiling of Tetraploid Astrocytes in Drug-Resistant Temporal Lobe Epilepsy

**DOI:** 10.64898/2026.01.30.26345206

**Authors:** Laura Cerrada-Gálvez, Rosario López-Rodríguez, Patricia Gonzalez-Tarno, Marcos Navares-Gómez, Paloma Pulido, Cristina Virginia Torres-Díaz, María C. Ovejero-Benito

**Affiliations:** Departamento de Ciencias Farmacéuticas y de la Salud, Facultad de Farmacia, Universidad San Pablo-CEU, CEU Universities, Urbanización Montepríncipe, 28660 Boadilla del Monte, Spain; Neurosurgery Department, Hospital Universitario de La Princesa, Madrid, Spain; Instituto de Investigación Sanitaria La Princesa (IIS-IP), Madrid, Spain; Clinical Pharmacology Department, Hospital Universitario de La Princesa, Faculty of Medicine, Universidad Autónoma de Madrid (UAM), Madrid, Spain

**Keywords:** astrocytes function, drug-resistant epilepsy, flow cytometry, polyploidy, reactive astrocytes, molecular pathogenicity

## Abstract

Epilepsy is one of the most prevalent neurological diseases, with 25-33% of patients developing drug-resistant epilepsy (DRE). The precise etiology of DRE remains unidentified. Recent studies have revealed an increase in tetraploid astrocytes in drug-resistant temporal lobe epilepsy (DR-TLE), a common subtype of DRE. This study aims to characterize the function of tetraploid astrocytes in the brain of subjects without central nervous system diseases and in DR-TLE. Cortical samples adjacent to the epileptogenic zone were obtained from DR-TLE patients undergoing resective neurosurgery and from postmortem donors without neurodegenerative, neurological, or psychiatric disorders. Tetraploid astrocytes were identified using the astrocytic marker NDRG2, and their functional characterization was assessed by evaluating markers of metabolism (ALDH1L1), transport (SOX9), electric function (NF1A), or reactive astrocytes (NFκB p65 and pSTAT3), via immunostaining followed by flow cytometry. Tetraploid astrocytes expressed all functional markers tested. The percentage of tetraploid astrocytes expressing ALDH1L1 or SOX9 was significantly increased in DR-TLE with respect to controls, whereas NF1A remained unchanged. Inflammatory markers pSTAT3 and NFκB p65 showed an upward trend in 4C astrocytes. In contrast, diploid (2C) astrocytes expressing these markers were reduced in DR-TLE, suggesting a functional shift toward polyploid cells in the DR-TLE cortex. Our findings suggest the preservation of markers of metabolism, transport and electric function in tetraploid astrocytes in physiological conditions and in DR-TLE patients. Moreover, the astrocytes with metabolic and transporter markers were significantly increased in DR-TLE. These findings point to tetraploid astrocytes as potential contributors to DR-TLE mechanisms.

## INTRODUCTION

Epilepsy is a chronic neurological disease characterized by the occurrence of spontaneous crises caused by exacerbated brain electrical activity [32]. Despite the development of new anti-seizure drugs (ASDs) [23], between 25-33% of patients do not show an adequate response [1, 33]. Drug-resistant epilepsy (DRE) is a frequent, costly, and debilitating disorder that presents a therapeutic challenge, significantly reducing patients’ quality of life and increasing mortality [26, 37]. This resistance to ASDs is especially prevalent in focal epilepsies such as temporal lobe epilepsy (TLE), one of the most common subtypes (drug-resistant TLE, DR-TLE).

The underlying mechanisms of DRE are not fully understood. Several hypotheses try to explain DRE, including drug-regulated or disease-intrinsic mechanisms, and pharmacokinetic or genetic factors [23]. Nevertheless, none fully explains the pathology [23]. Therefore, it is necessary to understand all the processes occurring in the brains of DRE patients to identify new therapeutic targets.

Tetraploid neurons, with a DNA content of 4C, have been described in different regions of the brain of subjects without central nervous system diseases where they perform physiological functions such as long-range projection [22]. Their numbers increase with aging and in neurodegenerative diseases such as Alzheimer’s [2, 20, 29], where they may play pathological roles [3]. Moreover, we recently reported a significant increase in tetraploid neurons in the epileptogenic zone of DRE patients [41].

Our group also described the presence of tetraploid astrocytes in different areas of the brain of subjects without central nervous system diseases and found that their proportion is significantly increased in the epileptogenic zone of DRE patients [41]. However, the functionality and contribution of tetraploid astrocytes to DR-TLE remain unknown.

Astrocytes are essential for maintaining physiological brain homeostasis [48, 49] by providing structural support, supplying energy to neurons through glucose metabolism, transporting and storing neurotransmitters, and modulating synaptic plasticity and electrical properties [7, 34, 39, 50]. Nevertheless, astrocytes play an active role in epileptogenesis by altering potassium and glutamate homeostasis, contributing to neuronal hyperexcitability and seizure propagation [11, 51]. In epilepsy, astrocytes may adopt a reactive phenotype, characterized by hypertrophy, increased proliferation [27, 38], and neuroinflammatory signaling, which contributes to the pathophysiology of DRE [48, 51].

The aim of this study is to characterize the function of tetraploid astrocytes in both physiological and pathological conditions such as DR-TLE.

## MATERIALS AND METHODS

### Study subjects and ethical approval

The protocol and the Informed Consent Form were approved by the Independent Clinical Research Ethics Committee of the *Hospital Universitario de La Princesa*. The revised Declaration of Helsinki and STROBE guidelines were followed to perform the study.

Samples of the surrounding cortex to the epileptogenic zone (temporal lobe) were obtained from DR-TLE patients and immediately frozen following neurosurgical resection. Samples were included in the collection registered in the Instituto de Salud Carlos III with the code C.0004381.

Postmortem cortical tissue was provided by the *Biobanco en Red de la Región de Murcia* (BIOBANC-MUR). Control samples were derived from donors without any history of neurodegenerative, neurological, or psychiatric diseases who had signed informed consents. These samples were examined by anatomopathologists to ensure the absence of pathological features. Furthermore, these samples were acquired and processed following standard operating procedures approved by their respective Ethics and Scientific Committees.

### Antibodies

The nuclear biomarker of adult astrocytes NDRG2 was chosen and detected using the rabbit polyclonal anti-NDRG2 antibody (Sigma-Aldrich, USA) diluted 1:500 to 1:55. Additionally, aldehyde dehydrogenase 1 family member L1 (ALDH1L1) was used as an indirect marker of metabolic function [52] and detected with the mouse monoclonal anti-ALDH1L1 antibody (sc-100497 Santa Cruz Biotechnology, USA), diluted 1:500. SRY-related high mobility group box gene 9 (SOX9) was employed to identify astrocytes with transport function [14, 45]. Sox9 was detected with the mouse monoclonal anti-SOX9 antibody (ab76997 Abcam, UK), diluted 1:500. Nuclear factor 1 (NF1A) was utilized as an indirect marker of electrical function [12], detected with the mouse monoclonal anti-NFA1 antibody (NBP3-13982 Bio-techne, USA) diluted 1:250. Finally, nuclear factor-κβ (NFκβ) p65 and p-STAT3 were used as reactive astrocytes biomarkers [14]. Mouse monoclonal anti-NFκβ p65 (12054-MM04 Sino Biological, USA) and anti-pSTAT3 (sc-8059 Santa Cruz Biotechnology, USA) antibodies were both diluted 1:500. To detect these primary antibodies, goat anti-rabbit DyLight 405 (A35350 Invitrogen, USA) diluted 1:500 and goat anti-mouse Alexa Fluor 647 (A21236 Invitrogen, USA), diluted 1:500, were used.

### Cell nuclei isolation and immunostaining

Using a dounce homogenizer with 3 mL of ice-cold homogenization buffer (PBS 0.1%Triton (PBT) with Pierce™ protease inhibitor tablets EDTA-free (Thermo Fisher Scientific, USA)) at ice-cold temperature to process each sample consisting of 3-5 mm edge-cubes of human cortical tissue. Cell nuclei isolation was carried out as described previously [21, 41].

Isolated unfixed cell nuclei immunostaining was conducted by adding primary and secondary antibodies at the same time within a medium comprising 10% Normal Goat Serum (NGS, Abcam, UK) and 5% bovine serum albumin (BSA, Sigma-Aldrich, USA) 1 mg/ml. A pool of all samples from each experiment was used to perform the controls: single stain controls and fluorescent minus one controls (FMOs. Subsequently, the reaction mixture was incubated overnight at 4ºC in darkness. Immunostained nuclei were filtered with a 40 μm nylon mesh (DDBiolab, Spain). The resulting volume was adjusted to 350–600 μl using DNase-free PBT buffer containing 50 μg/ml propidium iodide (PI, Sigma, USA) and 25 μg/ml DNase-free RNAse I (Sigma, USA). After incubating for 30 min at room temperature, the samples were analyzed in a Flow cytometer. DNA content was determined by PI incorporation and classified in three groups: diploid (2C), tetraploid (4C) and polyploid with a DNA content higher than 4C (>4C).

### Flow cytometry

Flow Cytometry was performed using an Attune NxT Acoustic Focusing Cytometer (Thermo Fisher Scientific Inc., USA) in *Instituto de Medicina Molecular Aplicada* (IMMA, Universidad San Pablo CEU). Data were analyzed with Thermo Scientific™ Attune™ NxT Software (Thermo Fisher Scientific, USA). Fluorophores DyLight 405, PI and Alexa Fluor 647 were excited with 405 nm, 488 nm or 637 nm laser and detected with VL1, BL2 and RL1 filters respectively. The gating strategy that was used in this study is described in Supplementary Figure 1.

### Statistical analysis

Statistical analyses were performed using R software, version 4.3.2 (http://www.R-project.org). Outliers were identified and removed using the interquartile range (IQR) method [10]. Potential batch effects were analyzed by applying the non-parametric Kruskal-Wallis test. Post hoc pairwise comparisons were assessed by corrected by Dunn’s test (corrected by Bonferroni). If a given experiment showed significant differences in seven or more of the twenty-eight gating parameters assessed, it was considered discordant and excluded from the analysis. Due to batch effect, one experiment was excluded from each of the SOX9, pSTAT3 and NFκβ p65 analyses. After data filtering, samples from at least 11 controls and 17 DR-TLE patients were analyzed for each marker.

Quantitative variables were represented by median and standard deviation. Statistical comparisons between DR-TLE patients and controls were performed using the Wilcoxon rank-sum test. Violin plots (combining boxplots with kernel density estimates) were used to visualize the expression of analyzed markers in the overall population and across 2C, 4C, and >4C astrocyte subpopulations. Demographic and clinical variables were shown as mean and standard deviations. A p-value < 0.05 was considered statistically significant.

## RESULTS

### Study population

A total of 35 samples were included in this study: 21 from DR-TLE patients with a mean age of 43.5 ± 11.6 years, and 14 from control subjects with a mean age of 60.1±10.9 years. The proportion of female participants was 20.6% in the DR-TLE group and 35.7% in the control group. Detailed clinical and demographic data are summarized in Table 1.

**Table 1.** Clinical and demographic characteristics of DR-TLE patients and control subjects included in this study. Data are shown as mean ± SD or number (percentage). Responders to neurosurgery were patients with an Engel value I and II. Abbreviations: DR-TLE, drug resistant temporal lobe epilepsy; NA, not applicable; NR, non-responders; R, responders.

|  |  | DR-TLE | Controls |
| --- | --- | --- | --- |
| Women |  | 7 (33.3) | 5 (35.7) |
| Age (years) | | 44 ± 12 | 60 ± 11 |
| Age at onset (years) | | 16 ± 16 | NA |
| Febrile seizures in childhood |  | 4 (19.0) | NA |
| Number of seizures/month | | 10 ± 16 | NA |
| Aura |  | 12 (57.1) | NA |
| Left dominant hand |  | 4 (19.0) | NA |
| Traumatism |  | 2 (9.5) | NA |
| Sensory, gait, or developmental disturbances |  | 4 (19.0) | NA |
| Neurosurgical hemisphere | Left | 13 (61.9) | NA |
|  | Right | 8 (38.1) | NA |
| Seizure etiology | Structural | 10 (47.6) | NA |
|  | Infectious | 2 (9.5) | NA |
|  | Unknown | 9 (42.9) | NA |
| Ever smoker |  | 8 (38.1) | NA |
| Alcohol |  | 6 (28.6) | NA |
| Responder Engel (12 months) |  | 12 (57.1) | NA |
| Responder Engel (24 months) |  | 8 (38.1) | NA |

### Astrocytes in DR-TLE

#### Overall

The overall percentage of polyploid astrocytes in the cortical tissue surrounding the epileptogenic zone was estimated by assessing the expression of the differentiated astrocytic marker NDRG2 using flow cytometry (Supplementary Figure 1). There were no significant differences in the percentage of NDRG2+ astrocytes between DR-TLE patients (n = 20) and controls (n = 14) (Supplementary Figure 2A). However, DR-TLE patients showed a significantly increased proportion of 4C astrocytes (p = 0.001) and >4C (p = 0.001) astrocytes compared to controls, indicating an enrichment of polyploid astrocytes in the cortex surrounding the epileptogenic zone (Supplementary Figures 2 B-D, Supplementary Table 1A).

#### Metabolic function

Given the critical role of astrocytes in cerebral metabolism, ALDH1L1 expression was evaluated to determine whether 4C astrocytes preserve this metabolic functional marker. The overall proportion of ALDH1L1-positive cells did not differ significantly between DR-TLE patients (n = 20) and controls (n = 14, Supplementary Figure 3A, Supplementary Table 1A), whereas stratification by DNA content revealed a significant reduction in 2C cells and an enrichment of 4C and >4C ALDH1L1-positive cells in DR-TLE patients (p <0.001, Supplementary Figures 3B-D, Supplementary Table1A).

The percentage of cells co-expressing NDRG2 and ALDH1L1 was also studied (Figure 1). No differences were observed in total NDRG2+ALDH1L1+ cells between controls and DR-TLE, whereas the percentage of NDRG2+ astrocytes expressing ALDH1L1 was significantly increased among 4C and >4C astrocytes in DR-TLE patients and decreased in 2C astrocytes (p <0.001) (Figure 1, Supplementary Table 1A).

**Figure 1.**
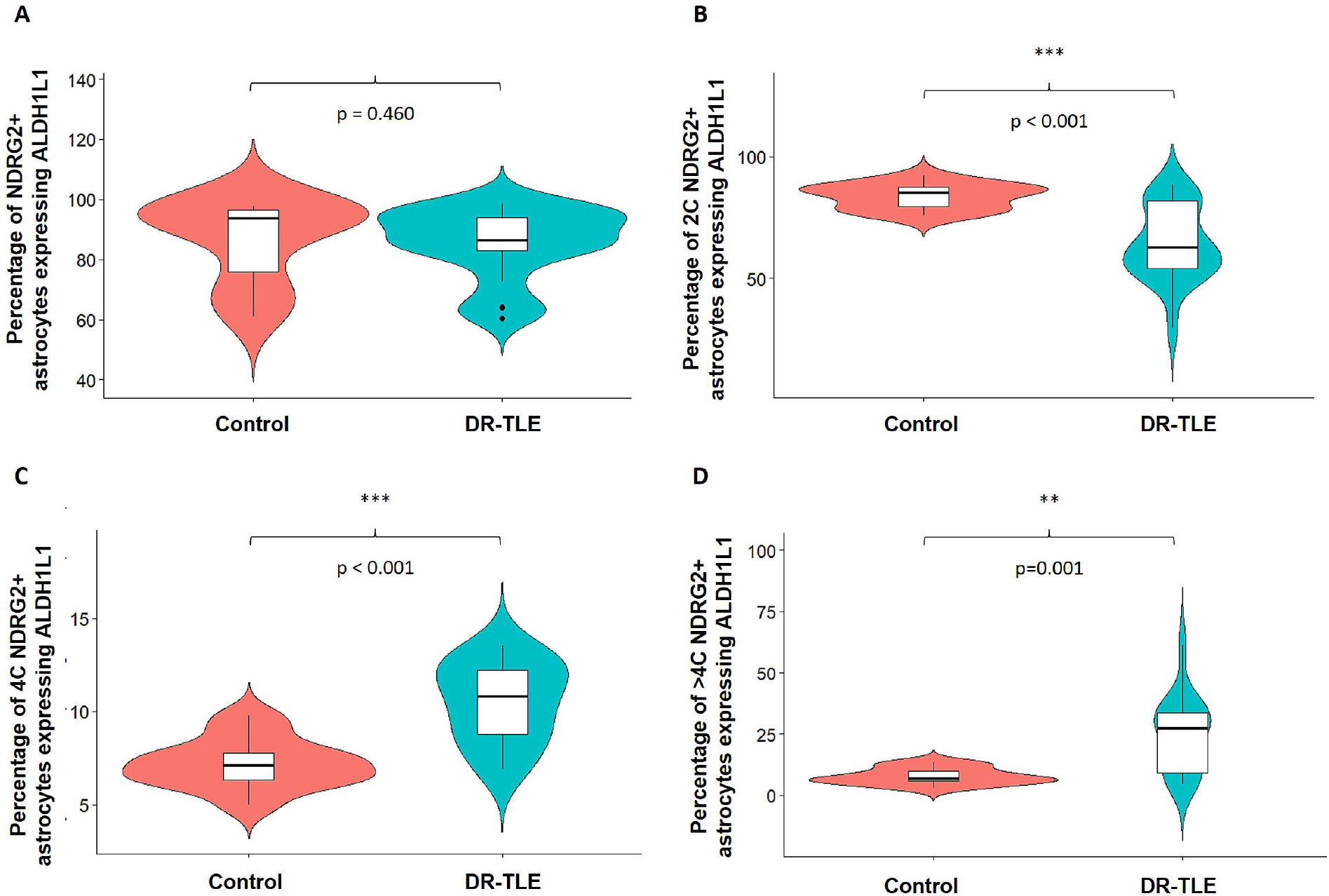
**A)** Violin box plot showing the percentage of (NDRG2+) expressing the metabolic biomarker ALDH1L1 in cortical tissue from controls (n = 14) and DR-TLE patients (n = 20). **B-D)** Percentage of astrocytes co-expressing NDRG2 and ALDH1L1 classified according to their DNA content. **B**) % 2C astrocytes; **C**) % 4C astrocytes; **D**) % >4C astrocytes. **p < 0.01, ***p < 0.001. Abbreviations: ALDH1L1: aldehyde dehydrogenase 1 family member L1; DR-TLE: drug-resistant temporal lobe epilepsy; NDRG2: N-Myc Downstream-Regulated Gene 2 Protein 2C: diploid; 4C: tetraploid; >4C: Polyploid with a DNA content higher than 4C.

#### Transport function

To assess potential alterations in transport-related astrocytic activity, the transcription factor SOX9 was examined. There were no significant differences in the overall proportion of SOX9-positive cells between DR-TLE patients (n = 18) and controls (n = 11, Supplementary Figure 4A, Supplementary Table 1B). However, DNA content analysis revealed a decrease in SOX9-positive cells (p <0.001) accompanied by a notable increase in 4C (p = 0.002) and >4C (p <0.001) SOX9-expressing nuclei (Supplementary Figures 4 B-D, Supplementary Table 1B).

Moreover, SOX9 and NDRG2 co-expression was also analyzed. In line with our findings, NDRG2+ SOX9+ expression was significantly enriched among 4C (p = 0.002) and >4C (p <0.001) astrocytes in DR-TLE patients and decreased in 2C astrocytes (p <0.001, Figure 2, Supplementary Table 1B).

**Figure 2.**
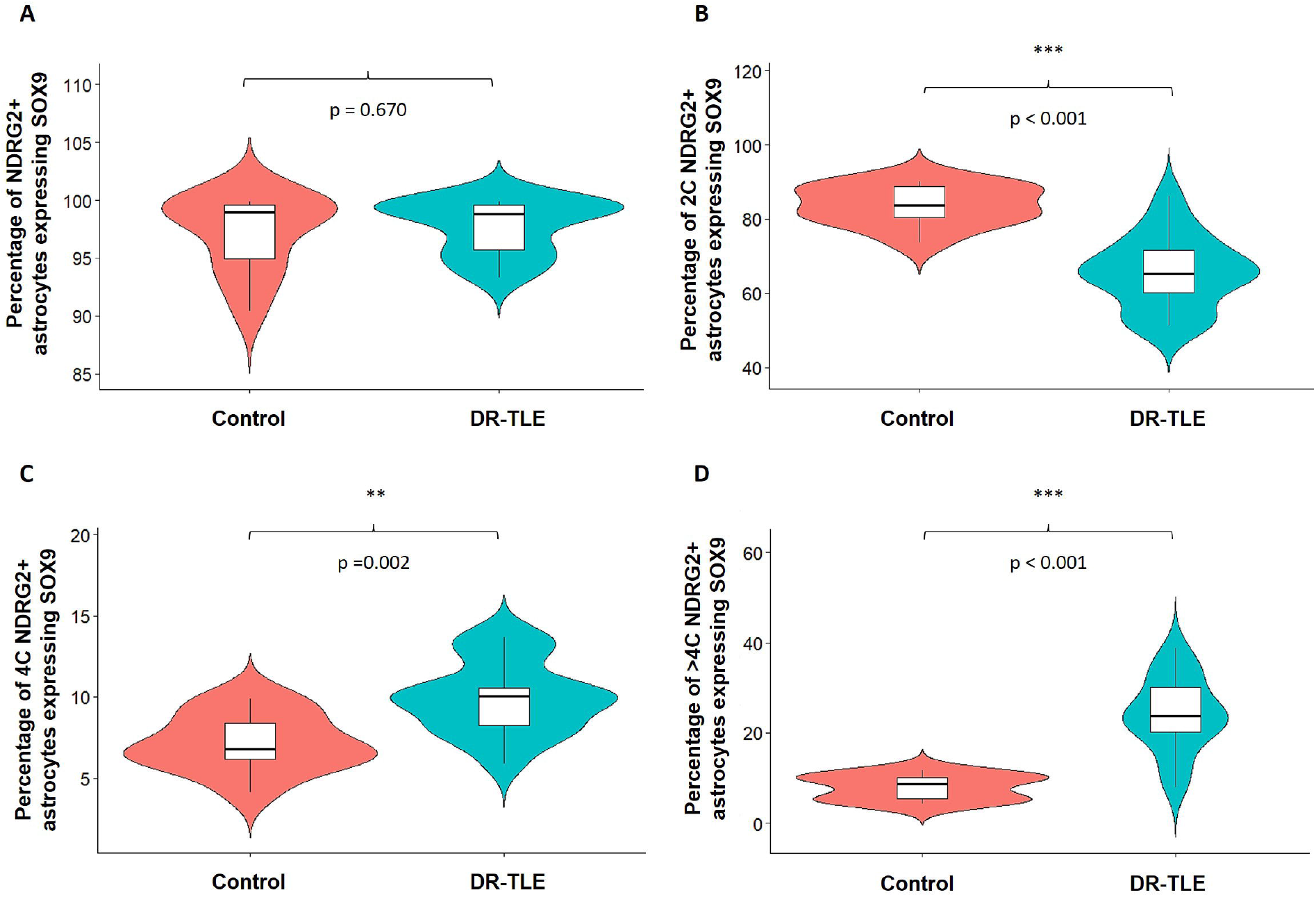
**A)** Violin box plot showing the percentage of astrocytes (NDRG2+) that express the transport biomarker SOX9 in cortical tissue from controls (n = 11) and DR-TLE patients (n = 18). **B-D**) Percentage of astrocytes co-expressing NDRG2 and SOX9 classified according to their DNA content. **B**) % 2C astrocytes; **C**) % 4C astrocytes; **D**) % >4C astrocytes. **p < 0.01, ***p < 0.001. Abbreviations: DR-TLE: drug-resistant temporal lobe epilepsy; NDRG2: N-Myc Downstream-Regulated Gene 2 Protein; SOX9: SRY-related high mobility group box gene 9; 2C: diploid; 4C: tetraploid; >4C: Polyploid with a DNA content higher than 4C.

#### Electrical function: NF1A

As part of their role in neuroglial regulation, astrocytes contribute to the modulation of neuronal excitability. The expression of NF1A was selected as a marker for astrocytic electrical function. The overall proportion of NF1A-positive cells did not differ significantly between patients (n = 17) and controls (n = 14, p = 0.190, Supplementary Figure 5 A, Supplementary Table 1C). However, a significant decrease in 2C NF1A-positive cells was observed (p <0.001), along with a proportional increase in 4C (p=0.021) and >4C (p <0.001) NF1A-positive cells within the DR-TLE group (Supplementary Figures 5 B-D, Supplementary Table 1C).

We analyzed double labeling of NF1A with NDRG2 to confirm that astrocytes express this electrical biomarker. Results confirmed a significant decrease in NF1A+/NDRG2+ 2C cells (p = 0.001) and an increase specifically among >4C astrocytes in DR-TLE patients (p <0.001), with no significant differences in the 4C population between groups (Figure 3, Supplementary Table 1C).

**Figure 3.**
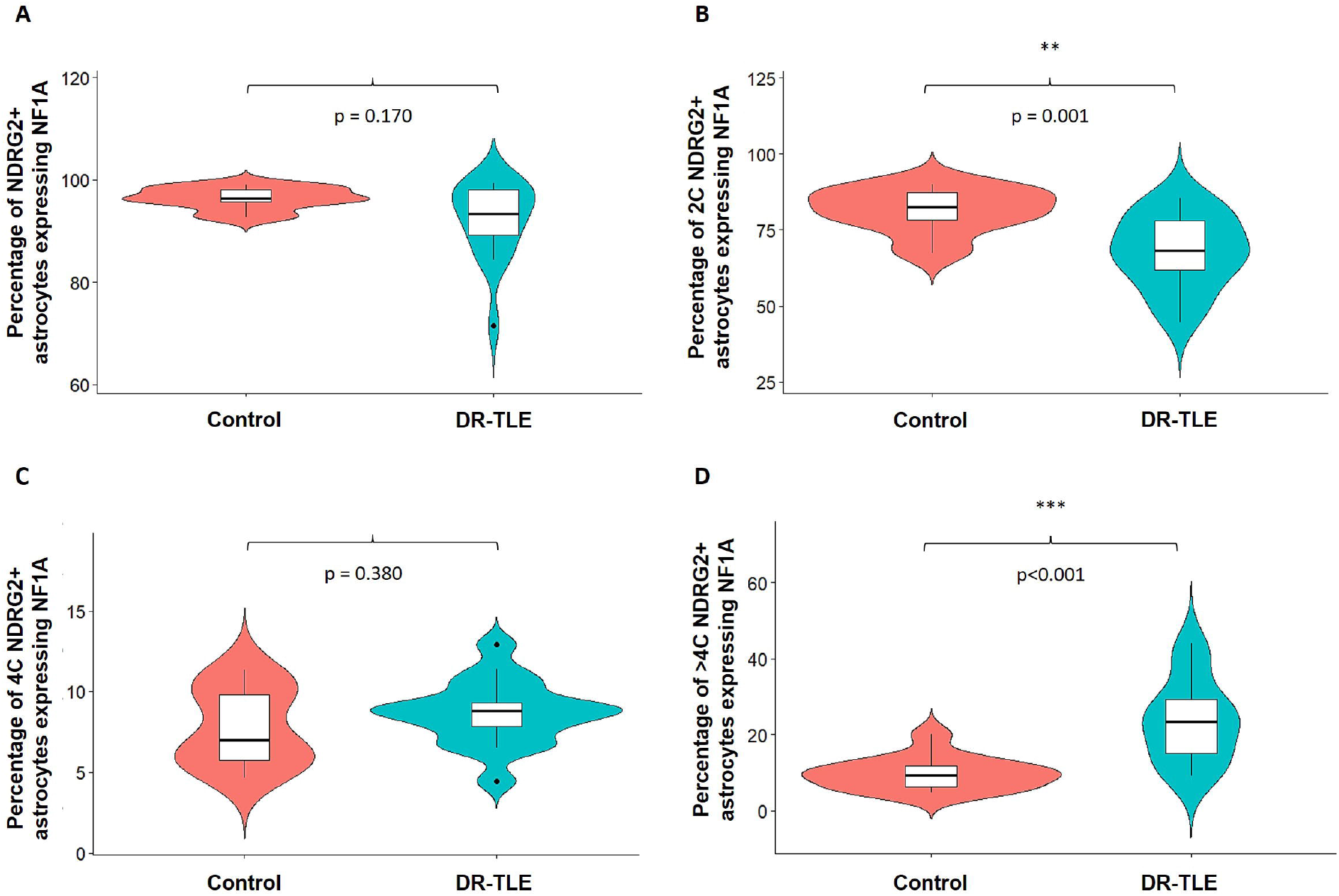
**A)** Violin box plot showing the percentage of astrocytes (NDRG2+) that express electrical biomarker NF1A in cortical tissue from controls (n = 14) and DR-TLE patients (n = 17). **B-D)** Percentage of astrocytes co-expressing NDRG2 and NF1A classified according to their DNA content. **B**) % 2C astrocytes; **C**) % 4C astrocytes; **D**) % >4C astrocytes. **p < 0.01, ***p < 0.001. Abbreviations: DR-TLE: drug-resistant temporal lobe epilepsy; NDRG2: N-Myc Downstream-Regulated Gene 2 Protein; NF1A: Nuclear factor 1; 2C: diploid; 4C: tetraploid; >4C: Polyploid with a DNA content higher than 4C.

#### Reactive astrocytes and inflammatory markers: pSTAT3 and NFκB p65

To investigate astrocyte-associated inflammatory signaling, the expression of two reactive astrocytes markers, pSTAT3 and NFκB p65, was analyzed. Although the overall percentage of pSTAT3+ and NFκB p65+ cells did not differ significantly between DR-TLE patients (n = 16-17) and controls (n = 12), stratification by ploidy revealed distinct alterations (Supplementary Figure 6, Supplementary Tables 1D and 1E). In DR-TLE samples, the proportion of 2C cells expressing either marker was significantly reduced (p <0.001). In contrast, NFκB p65+ 4C cells were significantly increased (p = 0.016, Supplementary Figure 6G, Supplementary Table 1E) while pSTAT3+ 4C cells showed an upward trend that did not reach statistical significance (p = 0.059, Supplementary Figure 6C; Supplementary Table 1D). The >4C cell populations exhibited a significant increase in expression of both markers (p <0.001, Supplementary Figures 6D and 6H Supplementary Tables 1D and 1E)

The total proportion of pSTAT3+/NDRG2+ and NFκB p65+/NDRG2+ astrocytes did not differ significantly between groups (Figures 4A and 4E). Similarly, the proportion of 4C astrocytes expressing these markers remained unchanged (Figures 4C and 4G). However, among 2C astrocytes in the patient group, expression of both pSTAT3+ (p < 0.001) and NFκB p65+ (p = 0.001) was significantly decreased, whereas the >4C population showed a marked increase (p <0.001) suggesting a redistribution of inflammatory signaling activity toward polyploid astrocytes in the DR-TLE brain (Figure 4, Supplementary Tables 1D and 1E).

**Figure 4.**
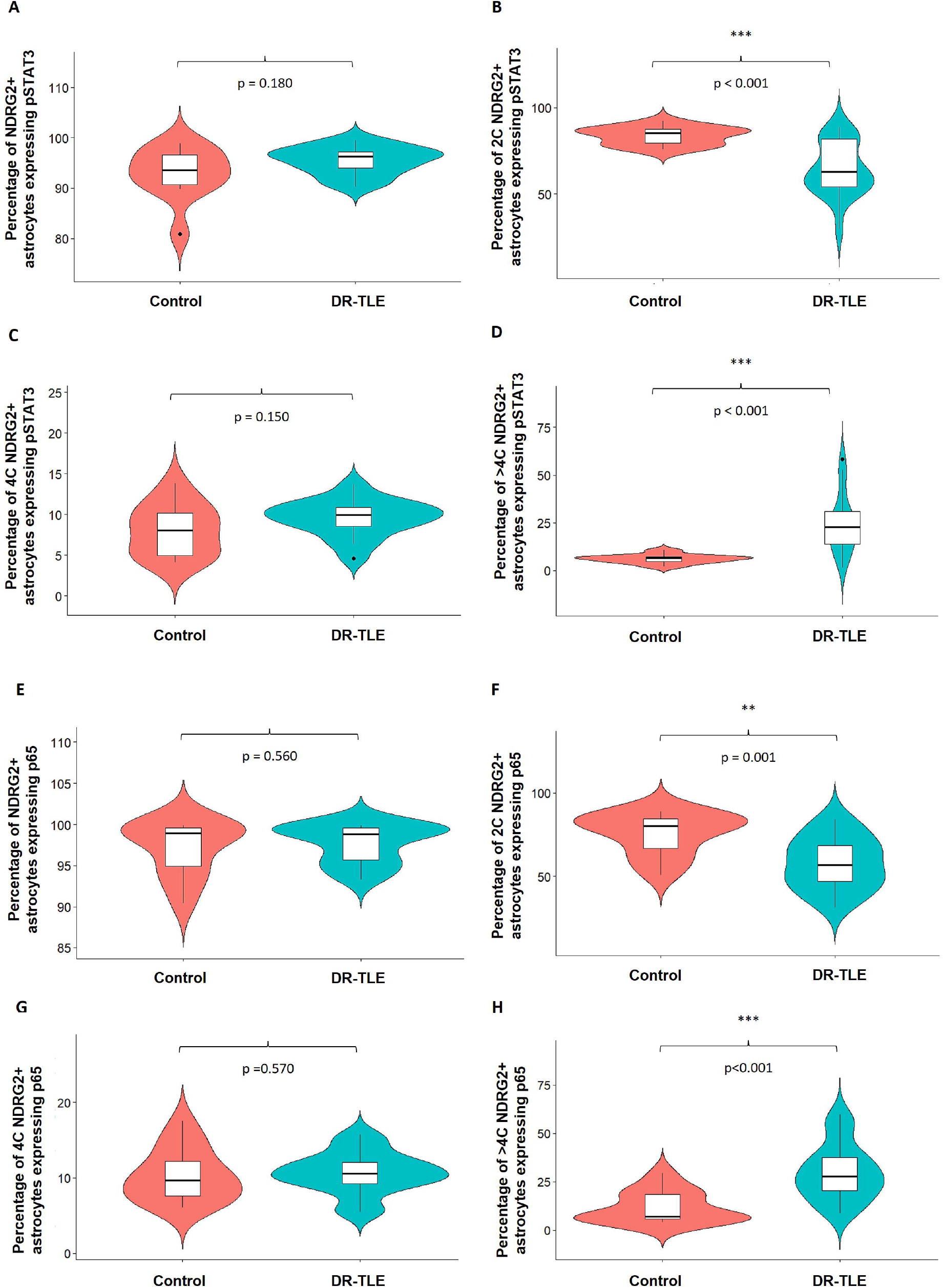
Violin box plot showing the percentage of differentiated astrocytes (NDRG2+) that express the reactive astrocytes biomarkers pSTAT3 (**A**) and NFκβ p65 (**E**) in cortical tissue from controls (n = 12) and DR-TLE patients (pSTAT3: n = 17; NFκβ p65: n = 16). **B-D)** Percentage of astrocytes co-expressing NDRG2 and pSTAT3 classified according to their DNA content. **B**) % 2C astrocytes; **C**) % 4C astrocytes; **D**) % >4C astrocytes. **F-H)** Percentage of astrocytes co-expressing NDRG2 and NFκβ p65 classified according to their DNA content. **F**) % 2C astrocytes; **G**) % 4C astrocytes; **H**) % >4C astrocytes. **p < 0.01, ***p < 0.001. Abbreviations: DR-TLE: drug-resistant temporal lobe epilepsy; NDRG2: N-Myc Downstream-Regulated Gene 2 Protein; p65: p65 subunit of nuclear factor-κβ; pSTAT3: phosphorylated Signal Transducer and Activator of Transcription 3; 2C: diploid; 4C: tetraploid; >4C: Polyploid with a DNA content higher than 4C.

## DISCUSSION

Our group previously described for the first time a higher presence of tetraploid astrocytes in DR-TLE patients compared to controls [41]. However, the functional characteristics of this subset of astrocytes remain unknown. In the present study, we observed that tetraploid astrocytes express biomarkers related to metabolic, transporter, and electrical function under both physiological conditions and DR-TLE (Figure 5). NDRG2 was used as an astrocyte marker because it labels a broad population of mature astrocytes [6, 8, 28], and is more widely expressed than GFAP, particularly in the cortex [54]. The overall percentage of astrocytes was similar in patients and controls, confirmed by the three biomarkers tested: NDRG2, ALDH1L1, and SOX9. Since not all astrocytes express NDRG2, additional markers were included in functional characterization.

**Figure 5.**
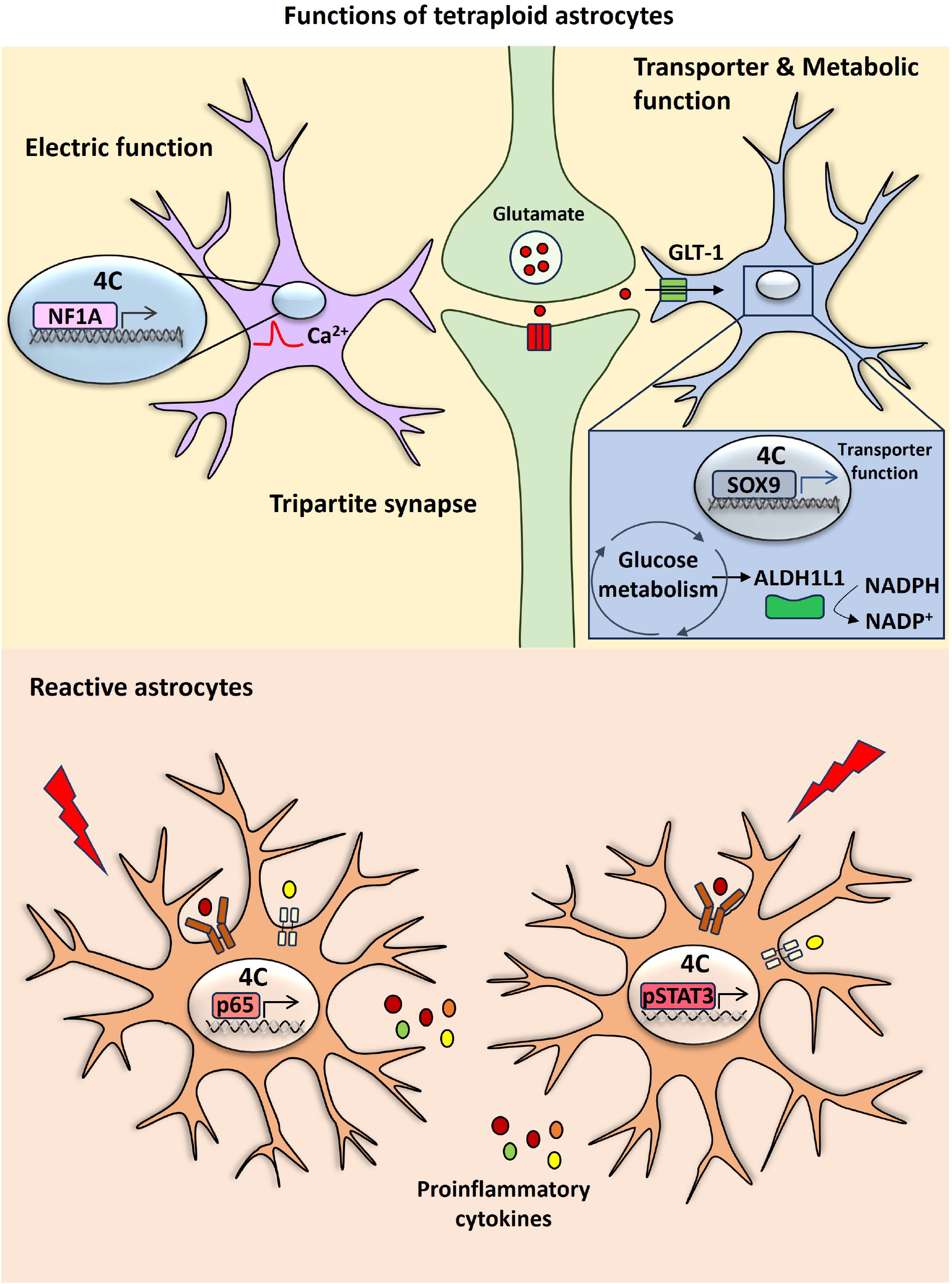
Diagram of the functions played by tetraploid astrocytes. Tetraploid astrocytes express NF1A suggesting that they are involved in electric function. Tetraploid astrocytes present SOX9, a marker of glutamate transporter function and ALDH1L1, an enzyme involved in glucose metabolism. Some of the tetraploid astrocytes have markers of reactive astrocytes (NFκB p65 and pSTAT3). Abbreviation: GLT-1: glutamate transporter 1; NADPH: Nicotinamide adenine dinucleotide phosphate; NDRG2: N-Myc Downstream-Regulated Gene 2 Protein; NF1A: Nuclear factor 1; p65: p65 subunit of nuclear factor-κβ; SOX9: SRY-related high mobility group box gene 9; pSTAT3: phosphorylated Signal Transducer and Activator of Transcription 3; 4C: tetraploid.

Astrocytic metabolic function was analyzed with ALDH1L1, which encodes the folate enzyme aldehyde dehydrogenase 1 family member L1, responsible for converting NADP to NADPH [14, 52]. ALDH1L1 is also expressed in radial glia [14] and neural stem cells [9], which may explain a higher percentage of ALDH1L1+ cells compared to NDRG2+/ALDH1L1+ cells in the cortex. This distribution was similar in controls and DR-TLE patients. Although one study conducted in an animal model of epilepsy did show exacerbated glucose metabolism during seizures, the metabolism and uptake of glucose decreased during the inter-ictal period [5]. The presence of ALDH1L1 marker in tetraploid astrocytes suggests preserved metabolic function, and the percentage was significantly increased in DR-TLE patients. The increase in tetraploid ALDH1L1+ astrocytes in DR-TLE patients, together with a proportional reduction of diploid ALDH1L1+ astrocytes, suggests a redistribution of metabolic capacity toward polyploid populations in DR-TLE.

SOX9 is a transcription factor involved in synapse function and immune reaction in astrocytes [31, 45]. SOX9-positive cells co-expresses most of the genes expressed by GLT1+ cells (glutamate transporter 1), suggesting that SOX9+ astrocytes can uptake glutamate, the excitatory neurotransmitter that has a key role in epilepsy [45]. We found that most tetraploid astrocytes co-express NDRG2 and SOX9, indicating preserved transport function in both the controls without central nervous system diseases and DR-TLE patients. Although a previous study in mice reported co-expression of GLT-1, SOX9, and ALDH1L1 in most cortical astrocytes [45], we observed a higher percentage of SOX9+ than ALDH1L1+ cells. Our results showed 30% of SOX9+ astrocytes, which is consistent with previous studies conducted in the human brain [18]. We did not observe differences in the percentage of astrocytes between DR-TLE patients and controls. However, the global ploidy distribution data indicates a general shift toward polyploidization in cortical astrocytes from DR-TLE patients. These results indicate a potential dysregulation in the astrocyte cell cycle, similar to the alterations previously reported in neuronal cells within the same pathological context [41].

Astrocytes modulate glutamate homeostasis by regulating the storage, metabolism and recycling of neurotransmitters [5]. Defective astrocyte glutamate uptake could contribute to the development of epilepsy [35]. In the intrahippocampal kainic acid (IHKA) model of TLE, GLT-1 immunoreactivity increases but then drops [13, 35]. Other publications reported a down-regulation of GLT-1 in animal models of epilepsy and proposed it as pharmacological target for epilepsy [43]. In line with these findings, pharmacological activation of GLT-1 may rescue learning and memory ability in the chronic phase of epileptogenesis [36]. We did not find significant differences in the percentage of astrocytes expressing SOX9 or ALDH1L1 between controls and DR-TLE patients, possibly due to analyzing the human cortex adjacent to the epileptogenic zone.

Astrocytes also participate in electrical transmission and tripartite synapse [31]. NF1A, a transcription factor expressed in oligodendrocytes, neurons and astrocytes, contributes to synaptic function and glial differentiation [12, 14, 17, 24]. Consistent with this, we observed a higher percentage of total NF1A+ cells compared to NDRG2+/NF1A+ astrocytes. NF1A promotes hippocampal astrocyte reactivity in a 4-AP-induced epileptic mouse model [16]. NF1A is upregulated in the epileptogenic zone of DRE patients which is related to the increase of pro-inflammatory cytokines levels [16]. However, we did not detect significant changes in the percentage of NF1A+ cells in the cortical region surrounding the epileptogenic zone. We found that most tetraploid astrocytes express NF1A, suggesting that their synaptic and electrical function remain stable, although there were no significant differences between controls and DR-TLE. Further additional electrophysiological studies should be carried out to confirm the electrical function of tetraploid astrocytes.

In recent years, there has been increasing evidence that neuroinflammation plays an important role in DRE [23, 25, 40]. Astrocytic dysfunction contributes to DRE by disrupting the blood-brain barrier, modulating drug transporters, and altering the inflammatory environment [49]. One of the key players in neuroinflammation is microglia, that releases pro-inflammatory cytokines which trigger astrocyte activation [15, 44]. Reactive astrocytes are characterized by increased proliferation, hypertrophy, cytokines release, and upregulation of markers such as pSTAT3, and NFκB p65 [27, 38]. NFκB is formed by two subunits (p65 and p50). Activation of surface receptors by proinflammatory cytokines triggers the transport of p65 into the nucleus where it works as a transcription factor of genes involved in processes such as cell survival genes or neuroinflammation [19]. In agreement with previous studies, we did not find differences in NFκB p65 levels between cortex surrounding the epileptogenic zone in DR-TLE and control samples [19]. Similarly, we did not find significant differences in the percentage of pSTAT3-positive cells between controls and DR-TLE, whereas pSTAT3 levels were increased in the hippocampus and the cortex of animal models of epilepsy [44]. Our data suggest that a subset of tetraploid astrocytes co-express both NFκB p65 and pSTAT3, suggesting a reactive phenotype that can be specific to 4C astrocytes under pathological conditions such as DR-TLE.

Interestingly, pSTAT3 promotes astrocytic cell cycle re-entry and DNA synthesis in epilepsy [42], possibly driving transformation from 2C to 4C and >4C astrocytes that remain functional [42]. These polyploid astrocytes may be a result of several rounds of endoreduplication (phases of synthesis without mitosis) [30]. Polyploid neurons with a DNA content higher than 4C were found in Alzheimer patients and are more prone to die by apoptosis than diploid neurons [2, 29]. In fact, hippocampal sclerosis, a hallmark of DR-TLE, features neuronal death and astrocyte proliferation [4, 46, 47]. As neuroinflammation is associated with cell cycle reactivation, it may promote polyploidy and subsequent apoptosis of neurons and astrocytes in DR-TLE. However, further studies should be carried out to confirm the connection between astrocytic tetraploidy and apoptosis.

It is important to highlight that this study was conducted using frozen cortical tissue from DR-TLE patients. While the sample size may appear modest, it is relatively high considering the difficulty of obtaining unfixed, human brain samples. The hippocampal area resected during neurosurgery is small and did not yield enough material to assess functional aspects of tetraploid astrocytes. Another limitation in our study is the absence of postmortem brain samples from epilepsy patients responding to ASDs. Furthermore, the use of frozen tissue may compromise cellular integrity, which prevented us from using classical membrane biomarkers of astrocytic function such as GABA, AQP4 or NMDA [39, 48, 53]. Nevertheless, the protocol used to detect the nuclei of tetraploid cells has been validated in previous studies [20, 22, 41]. An additional advantage of targeting nuclear markers is that nuclei are generally more resistant to the disruption caused by tissue dissociation of the whole cells [45].

## CONCLUSION

To our knowledge, this study provides the first functional characterization of tetraploid astrocytes in the human brain under both physiological and pathological conditions such as DR-TLE. These findings indicate that tetraploid astrocytes have functional capacities, including metabolic, transporter, and electrical functions, similar to diploid astrocytes. In DR-TLE patients, there is a significant increase in 4C and >4C astrocytes, accompanied by enhanced expression of these with the metabolic (ALDH1L1) and transporter (SOX9) markers. Although electrical function (NF1A) remained unchanged, a significant redistribution of inflammatory markers (pSTAT3 and NFκB p65) toward polyploid astrocytes was observed in DR-TLE patients, suggesting that these cells may participate in neuroinflammatory processes. Moreover, the decrease in the percentage of 2C astrocytes expressing these markers in DR-TLE patients supports the hypothesis that these cells convert to functional polyploid cells in the cortex surrounding the epileptogenic zone. Our results suggest that 4C astrocytes may contribute to altered homeostatic and inflammatory microenvironment of DR-TLE.

Further studies are needed to elucidate the mechanisms underlying astrocytic tetraploidization and its potential role in epileptogenesis and disease progression. However, these results open new avenues for understanding astrocyte roles in epilepsy and in the future may identify new therapeutic targets.

## Supplementary Tables

**Supplementary Table 1:** Percentage of cells within each flow-cytometry gate for control and DR-TLE patient groups. Flow cytometry gates are detailed in Supplementary Figure 1. **A)** ALDH1L1 (controls n = 14; DR-TLE n = 20); **B)** SOX9 (controls n = 11; DR-TLE n = 18); **C)** NF1A (controls n = 14; DR-TLE n = 17); **D)** pSTAT3 (controls n = 12; DR-TLE n = 16); **E)** p65 (controls n = 14; DR-TLE n = 18). Values are shown as median and group comparisons were performed with the Wilcoxon rank-sum test. Abbreviations: ALDH1L1: aldehyde dehydrogenase 1 family member L1; DR-TLE: drug-resistant temporal lobe epilepsy; Min: minimum; Max: Maximum; NF1A: Nuclear factor 1; NDRG2: N-Myc Downstream-Regulated Gene 2 Protein; p65: p65 subunit of nuclear factor-κβ; PI: propidium iodide; Sox9: SRY-related high mobility group box gene 9; pSTAT3: phosphorylated Signal Transducer and Activator of Transcription 3; 2C: diploid; 4C: tetraploid; >4C: Polyploid with a DNA content higher than 4C.

## Supplementary Figures

**Supplementary Figure 1**. Gating strategy for the identification of tetraploid nuclei by flow cytometry. Histograms are derived from a representative DR-TLE patient. The color of the square surrounding each plot indicates the parent gate from which they are derived. **A)** Nuclei were gated based on size and granularity using forward scatter area (FSC-A) and side scatter area (SSC-A) parameters. **B)** Nuclei were discriminated against debris by their ability to bind to PI. **C)** DNA content was analyzed by plotting PI-Height (BL2-A, PI-H) against PI-Area (BL2-A, PI-A) within the gated population. PI was detected using the BL2 filter. Singlets PI+ population is highlighted in dark green. **D)** Overlay histogram showing specific SOX9+ staining detected with the RL1 filter in the singlets population. The black square shows the SOX9+ population. **E)** SOX9+ cells classified in 2C, 4C and >4C according to their PI incorporation. **F)** Overlay histogram showing specific NDRG2+ staining detected with the VL1 filter in the PI+ singlets population. NDRG2+ population is highlighted with a blue square. **G)** NDRG2+ cells classified in 2C, 4C and >4C according to their PI incorporation. **J)** Histogram showing singlets PI+ NDRG2+ co-expressing SOX9 from the NDRG2+ population (Supplementary Figure 1F). The purple square highlights cells co-expressing NDRG2+ and SOX9+. **K)** Histogram showing cells co-expressing NDRG2 and SOX9+ classified in 2C, 4C and >4C according to their DNA content. **H)** Histogram showing DNA content of singlets PI+ represented in dark green (Supplementary Figure 1C) classified according to their DNA content in 2C, 4C and >4C cells. The red, yellow and brown squares represent the 2C, 4C and >4C populations, respectively. **I)** Histogram showing the percentage of 2C cells expressing SOX9 from the 2C population. **L)** Histogram showing the percentage of 2C cells expressing NDRG2 from the 2C population, highlighted in pale green. **M)** Histogram showing the percentage of 2C cells co-expressing NDRG2 and SOX9+ from the 2C NDRG2+ population (represented in pale green). **N)** Histogram showing the percentage of 4C cells expressing SOX9 from the 4C population. **O)** Histogram showing the percentage of 4C cells expressing NDRG2 from the 4C population, highlighted in grey. **P)** Histogram showing the percentage of 4C cells co-expressing NDRG2 and SOX9+ from the 4C NDRG2+ population (represented in grey). **Q)** Histogram showing the percentage of >4C cells expressing SOX9 from the >4C population. **R)** Histogram showing the percentage of >4C cells expressing NDRG2 from the >4C population, highlighted in pink. **S)** Histogram showing the percentage of >4C cells co-expressing NDRG2 and SOX9+ from the >4C NDRG2+ population (represented in pink). SOX9 was used as an example of the different markers. This strategy was repeated with the different markers of astrocyte function: ALDH1L1, NF1A, pSTAT3 and NFκβ p65. Abbreviations: ALDH1L1: aldehyde dehydrogenase 1 family member L1; BL2: Blue light 2 filter; DR-TLE: drug-resistant temporal lobe epilepsy; NF1A: Nuclear factor 1; NDRG2: N-Myc Downstream-Regulated Gene 2 Protein; p65: p65 subunit of nuclear factor-κβ; PI: propidium iodide; RL1: Red light 1 filter; SOX9: SRY-related high mobility group box gene 9; pSTAT3: phosphorylated Signal Transducer and Activator of Transcription 3; VL1: Violet light 1 filter; 2C: diploid; 4C: tetraploid; >4C: Polyploid with a DNA content higher than 4C.

**Supplementary Figure 2. A)** Violin box plot showing the percentage of total differentiated astrocytes (NDRG2+ cells) in the cortical tissue from controls (n = 14) and patients with DR-TLE (n = 21). **B-D)** Percentage of astrocytes expressing classified according to their DNA content. **B)** 2C astrocytes; **C)** 4C astrocytes; **D)** >4C astrocytes. *p < 0.05, **p < 0.01, ***p < 0.001. Abbreviations: DR-TLE: drug-resistant temporal lobe epilepsy; NDRG2: N-Myc Downstream-Regulated Gene 2 Protein; 2C: diploid; 4C: tetraploid; >4C: Polyploid with a DNA content higher than 4C; ns: no significant differences.

**Supplementary Figure 3. A)** Violin box plot showing the percentage of total ALDH1L1-positive cells in the cortical tissue from controls (n = 14) and patients with DR-TLE (n = 20). **B-D)** Percentage of ALDH1L1-positive cells expressing classified according to their DNA content. **B)** 2C astrocytes; **C)** 4C astrocytes; **D)** >4C astrocytes. *p < 0.05, **p < 0.01, ***p < 0.001. Abbreviations: ALDH1L1: aldehyde dehydrogenase 1 family member L1; DR-TLE: drug-resistant temporal lobe epilepsy; 2C: diploid; 4C: tetraploid; >4C: Polyploid with a DNA content higher than 4C; ns: no significant differences.

**Supplementary Figure 4. A)** Violin box plot showing the percentage of total SOX9-positive cells in the cortical tissue from controls (n = 11) and patients with DR-TLE (n = 18). **B-D)** Percentage of SOX9-positive cells expressing classified according to their DNA content. **B)** 2C astrocytes; **C)** 4C astrocytes; **D)** >4C astrocytes. *p < 0.05, **p < 0.01, ***p < 0.001. Abbreviations: DR-TLE: drug-resistant temporal lobe epilepsy; Sox9: SRY-related high mobility group box gene 9; 2C: diploid; 4C: tetraploid; >4C: Polyploid with a DNA content higher than 4C; ns: no significant differences.

**Supplementary Figure 5. A)** Violin box plot showing the percentage of total NF1A-positive cells in the cortical tissue from controls (n = 14) and patients with DR-TLE (n = 17). **B-D)** Percentage of cells expressing NF1A classified according to their DNA content. **B**) % 2C astrocytes; **C**) % 4C astrocytes; **D**) % >4C astrocytes. *p < 0.05, **p < 0.01, ***p < 0.001. Abbreviations: DR-TLE: drug-resistant temporal lobe epilepsy; NF1A: Nuclear factor 1; 2C: diploid; 4C: tetraploid; >4C: Polyploid with a DNA content higher than 4C; ns: no significant differences.

**Supplementary Figure 6**. A) Violin box plot showing the percentage of total pSTAT3-positive cells in the cortical tissue from controls (n = 12) and patients with DR-TLE (n = 17). **B-D)** Percentage of cells expressing pSTAT3 classified according to their DNA content. **B**) % 2C astrocytes; **C**) % 4C astrocytes; **D**) % >4C astrocytes. E) Violin box plot showing the percentage of total NFκβ p65-positive cells in the cortical tissue from controls (n = 14) and patients with DR-TLE (n = 20). **F-H)** Percentage of cells expressing NFκβ p65 classified according to their DNA content. **F**) % 2C astrocytes; **G**) % 4C astrocytes; **H**) % >4C astrocytes. *p < 0.05, **p < 0.01, ***p < 0.001. Abbreviations: DR-TLE: drug-resistant temporal lobe epilepsy; p65: p65 subunit of nuclear factor-κβ; pSTAT3: phosphorylated Signal Transducer and Activator of Transcription 3; 2C: diploid; 4C: tetraploid; >4C: Polyploid with a DNA content higher than 4C.

## ACKNOWLEDGEMENTS

We thank the epilepsy patients who kindly donated brain samples. We are particularly grateful for the generous contribution of the patients and the collaboration of the Biobank Network of the Region of Murcia, BIOBANC-MUR, registered on the Registro Nacional de Biobancos with registration number B.0000859. BIOBANC-MUR is supported by the “Instituto de Salud Carlos III” (proyecto PT20/00109), by “Instituto Murciano de Investigación Biosanitaria Virgen de la Arrixaca, IMIB” and by “Consejería de Salud de la Comunidad Autónoma de la Región de Murcia”. We would like to thank Gillian Moody for her help with the study and their valuable comments on this manuscript. We also thank our colleagues from the Flow Cytometry Unit of “Instituto de Medicina Molecular Aplicada” (IMMA, Universidad San Pablo CEU).

This study was supported by Instituto de Salud Carlos III: PI20/01391. Laura Cerrada-Gálvez has a grant awarded by the Fundación Universitaria San Pablo CEU and Banco Santander “Ayudas a la Formación de Jóvenes Investigadores CEU-Santander”.

## DECLARATION OF INTERESTS

The authors have no relevant financial or non-financial interests to disclose.

## DATA AVAILABILITY

The data that support the findings of this study are available from the corresponding author upon reasonable request.

## CRedIT Author contributions

LCG: Investigation, Formal analysis, Data curation, Methodology, Visualization and Writing; RLR: Formal analysis and Data curation; PP, MNG: Resources; CVT: Resources, Project administration and Funding acquisition; MCOB: Conceptualization, Formal analysis, Methodology, Investigation, Visualization, Validation, Project administration, Funding acquisition, Supervision and Writing - Original Draft. All authors have read, reviewed, and approved the final manuscript.

## Notes

### Competing Interest Statement

The authors have declared no competing interest.

### Funding Statement

This study was supported by Instituto de Salud Carlos III: PI20/01391. Laura Cerrada-Galvez has a grant awarded by the Fundacion Universitaria San Pablo CEU and Banco Santander Ayudas a la Formacion de Jovenes Investigadores CEU- Santander.

### Author Declarations

The protocol and the Informed Consent Form were approved by the Independent Clinical Research Ethics Committee of the Hospital Universitario de La Princesa. The revised Declaration of Helsinki and STROBE guidelines were followed to perform the study.

