## Supplementary figures and images for "Functional Profiling of Tetraploid Astrocytes in Drug-Resistant Temporal Lobe Epilepsy"

### Supplementary Figure 1

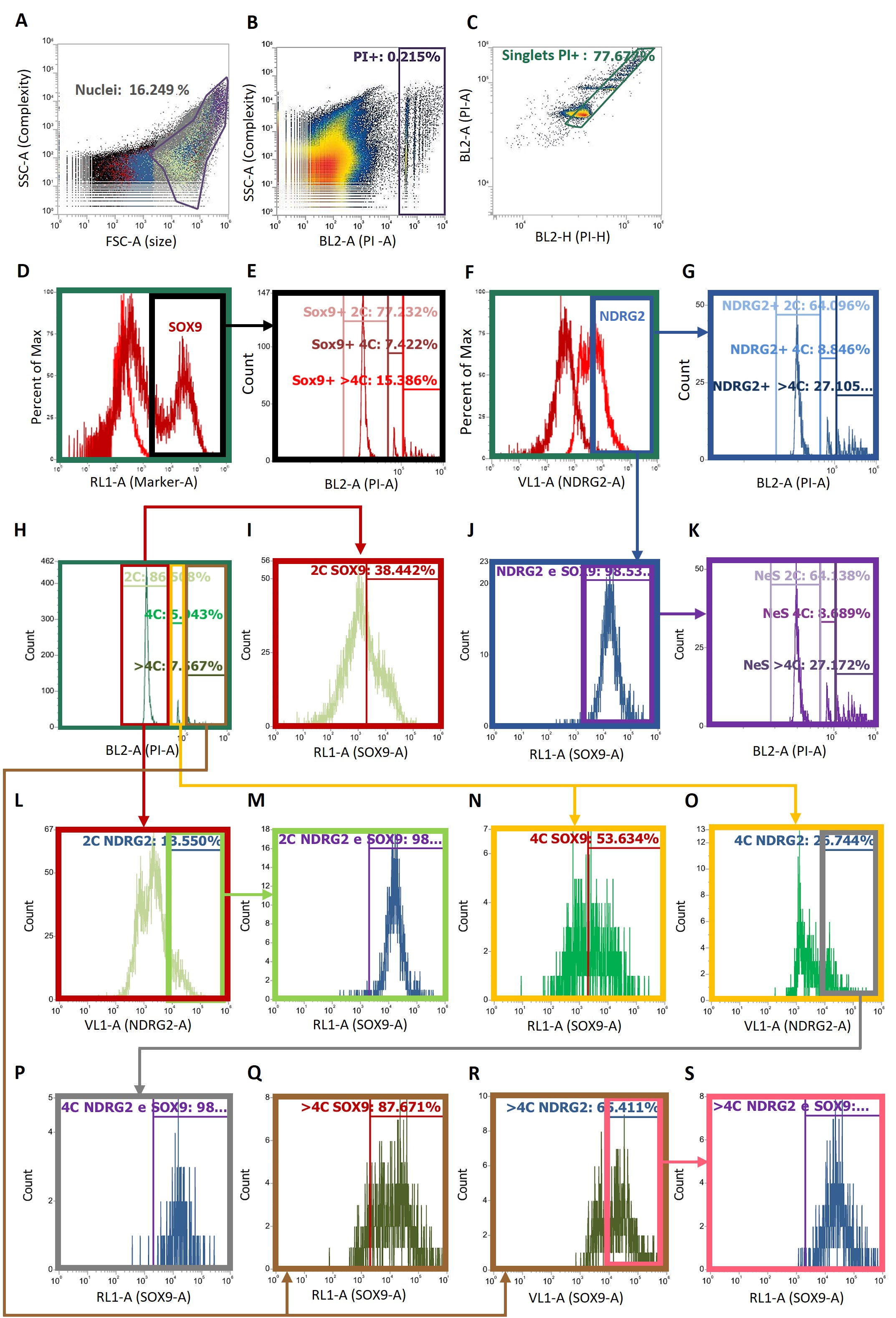

### Supplementary Figure 2

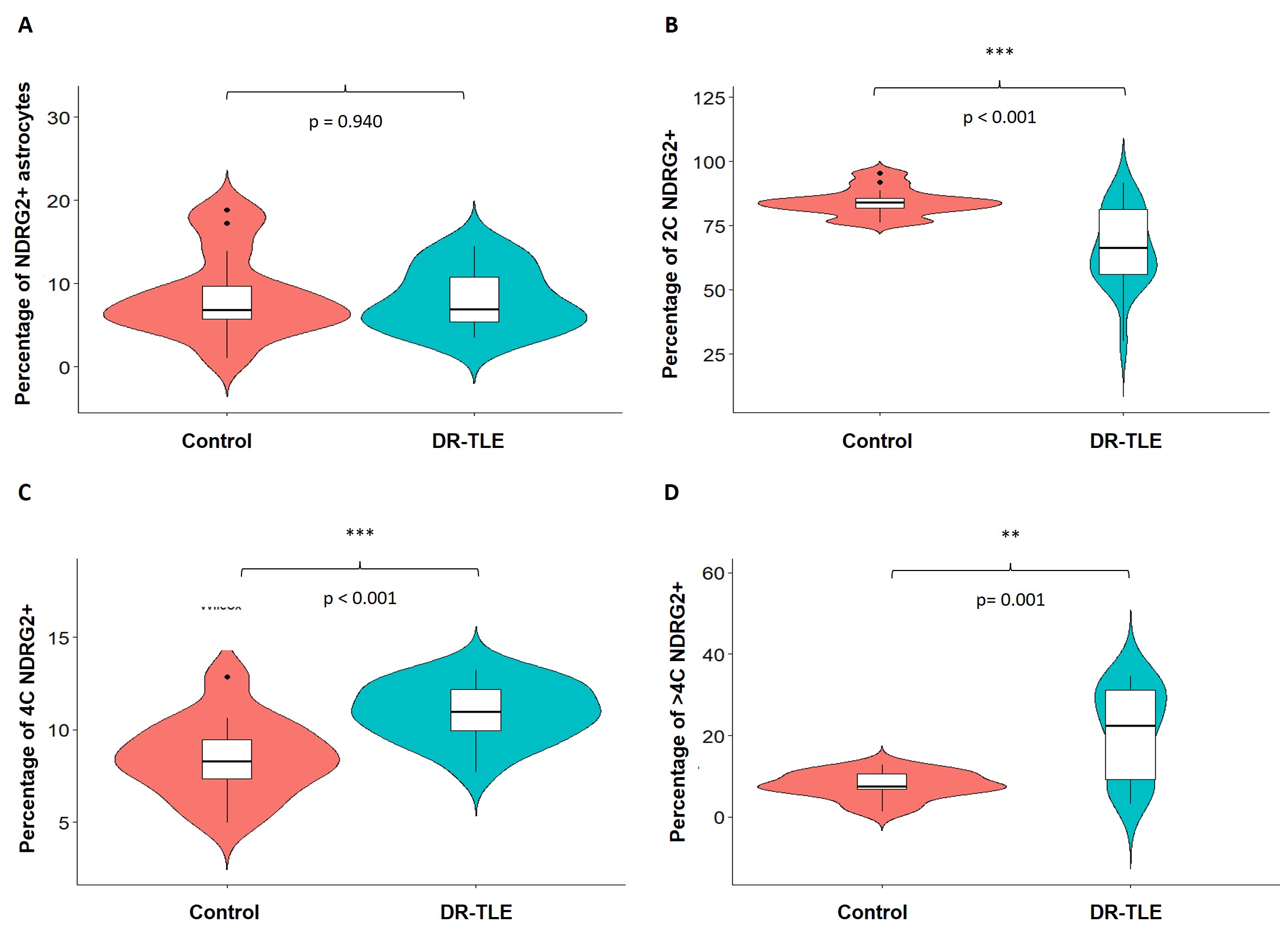

### Supplementary Figure 3

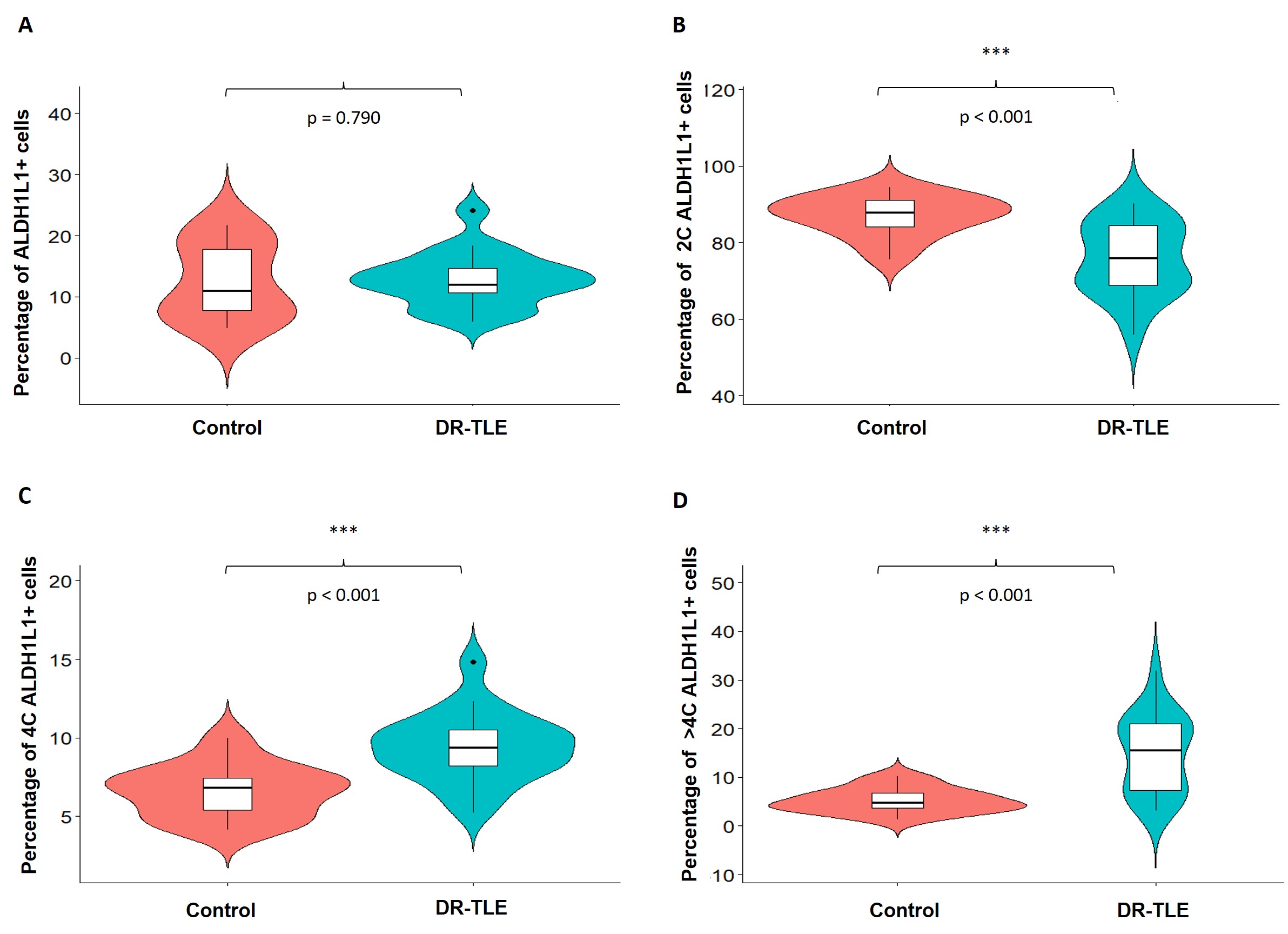

### Supplementary Figure 4

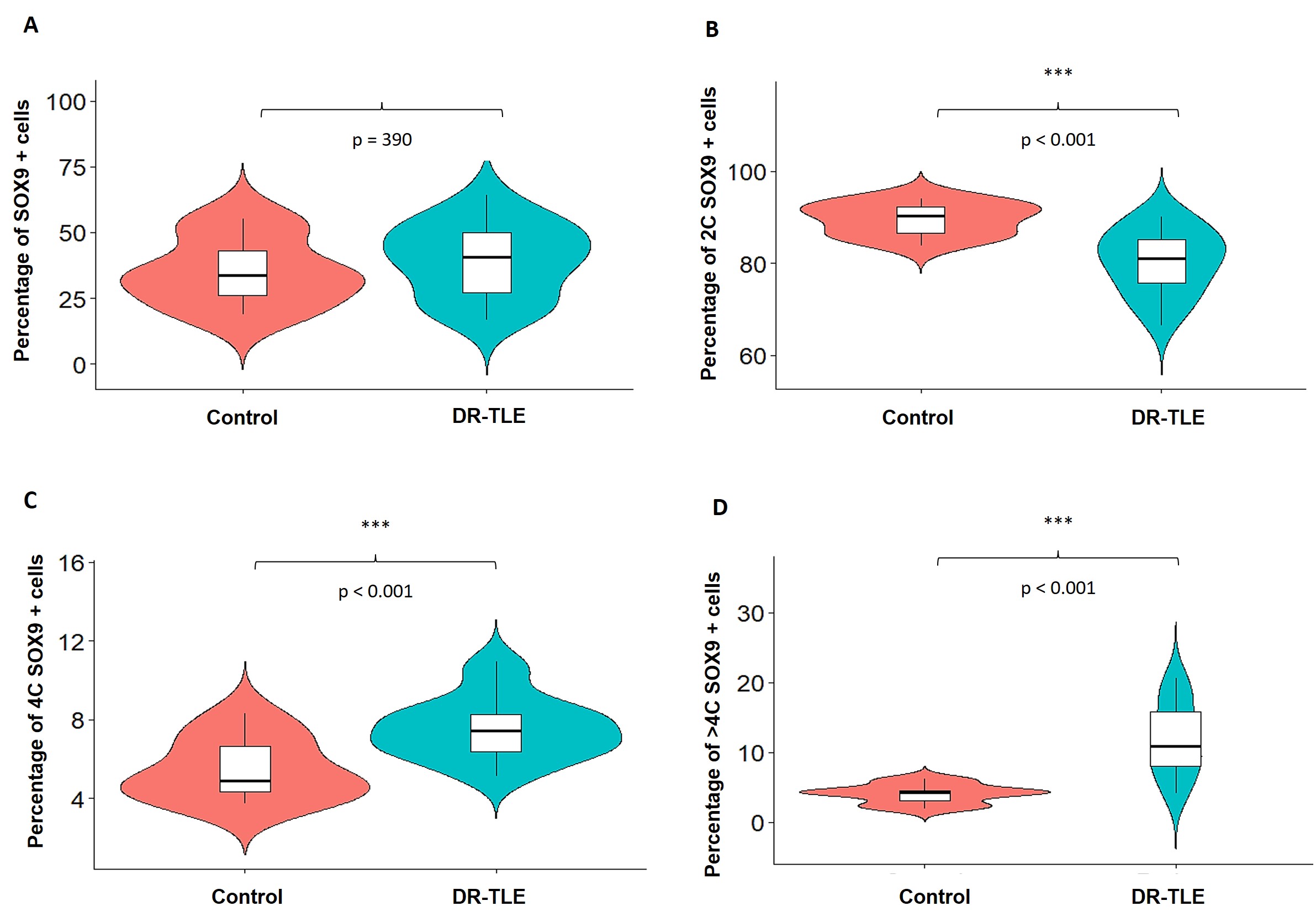

### Supplementary Figure 5

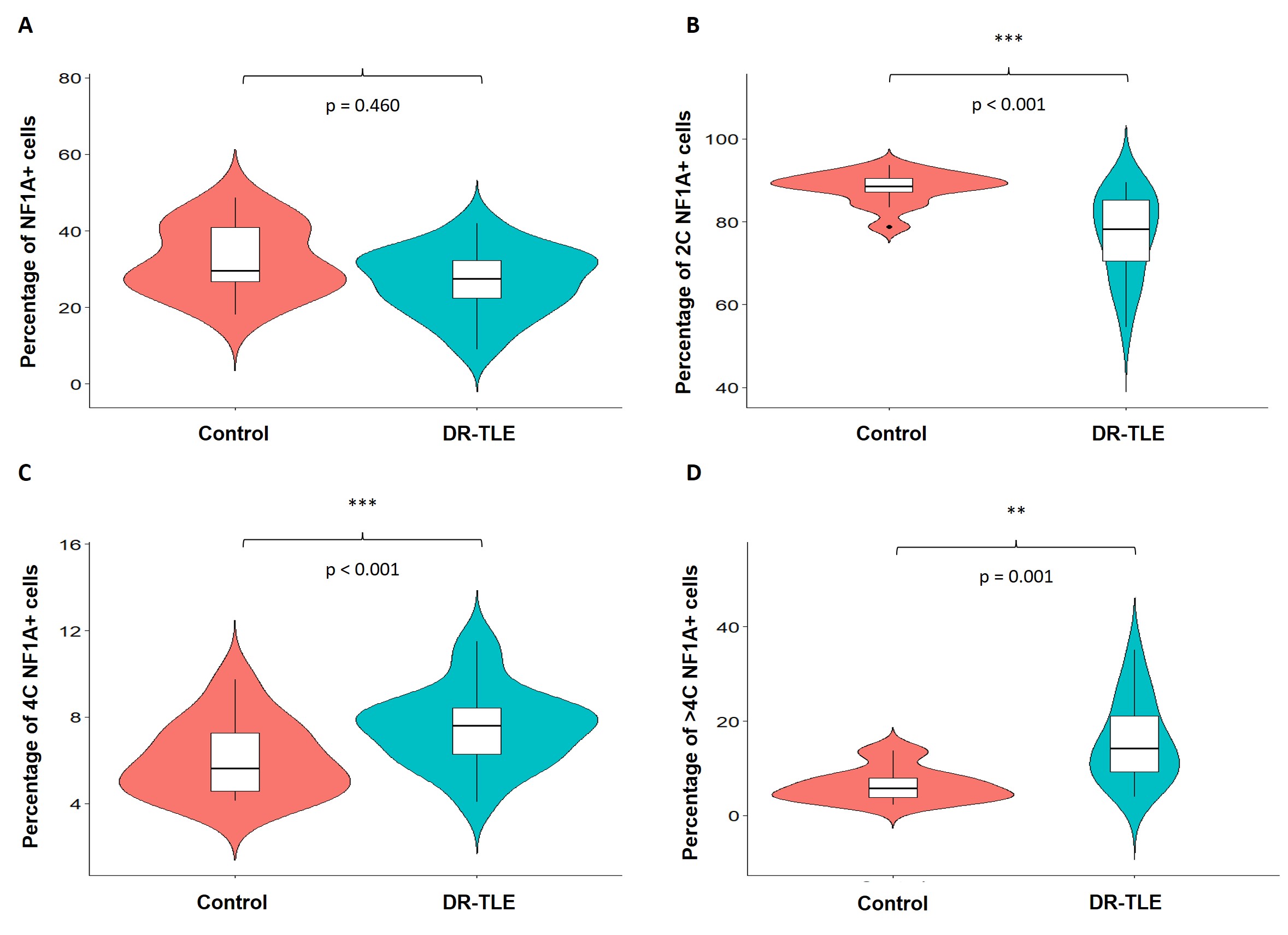

### Supplementary Figure 6

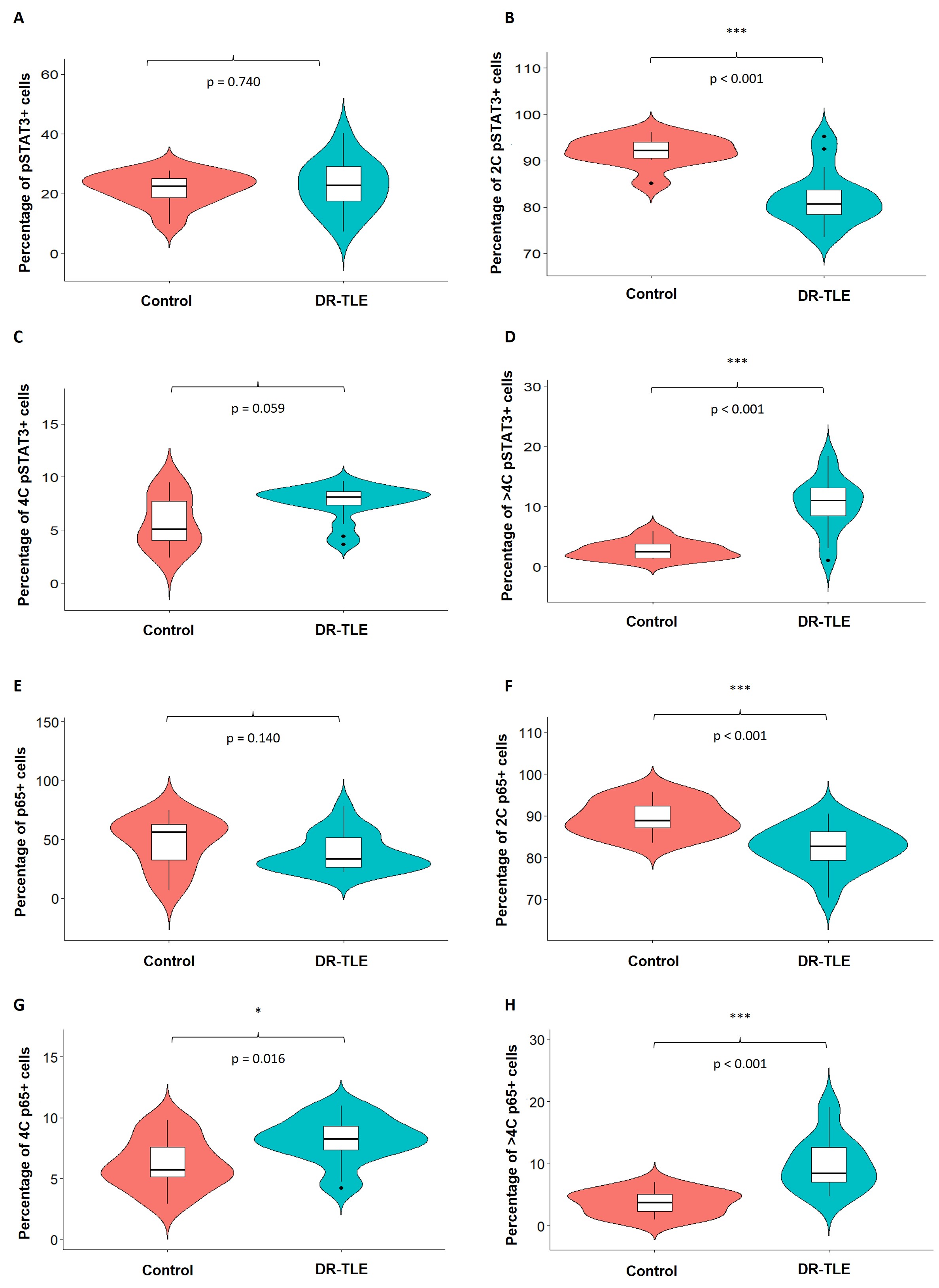
